# Modelling for prediction of the spread and severity of COVID-19 and its association with socioeconomic factors and virus types

**DOI:** 10.1101/2020.06.18.20134874

**Authors:** Shreshth Tuli, Shikhar Tuli, Ruchi Verma, Rakesh Tuli

## Abstract

We report the development of a Weibull based Long-Short-Term-Memory approach (W-LSTM) for the prediction of COVID-19 disease. The W-LSTM model developed in this study, performs better in terms of MSE, *R*^2^ and MAPE, as compared to the previously published models, including ARIMA, LSTM and their variations. Using W-LSTM model, we have predicted the beginning and end of the current cycle of COVID-19 in several countries. Performance of the model was validated as satisfactory in 82% of the 50 test countries, while asking for prediction for 10 days beyond the period of training. Accuracy of the above prediction with days beyond training was assessed in comparison with the MAPE that the model gave with cumulative global data. The model was applied to study correlation between the growth of infection and deaths, and a number of effectors that may influence the epidemic. The model identified age groups, trade with China, air traffic, country temperature and CoV-2 virus types as the likely effectors of infection and virulence leading to deaths. The predictors likely to promote or suppress the epidemic were identified. Some of the predictors had significant effect on the shape parameters of Weibull distribution. The model can function on cloud, take inputs in real time and handle large data country wise, at low costs to make predictions dynamically. Such predictions are highly valuable in guiding policy makers, administration and health. Interactive curves generated from the W-LSTM model can be seen at http://collaboration.coraltele.com/covid2/.

## 1. Introduction

Since the first few reports from Wuhan, China in December, 2019, the Coronavirus disease COVID-19 has now spread globally to more than 200 countries and territories. It has been declared a pandemic by WHO. In the absence of a curative drug or vaccine, containing the disease by non-pharmaceutical approaches is the highest priority to guard against its continued outbreak and spread. It is caused by a member of the SARS group of viruses, called novel Coronavirus CoV-2, which is more contagious than the previously known SARS viruses. In a large proportion of the cases, CoV-2 incubates in individuals with no exhibited symptoms, who continue to spread the infection. Within a period of less than 6 months, more than six million people have globally been tested as infected by CoV-2, which has claimed some 370000 lives. Containing and managing the spread of COVID-19 requires anticipating well in advance, the magnitude and dimension that a pandemic like this, may take in coming months.

Mathematical models for simulating the phase based human transmissibility of the disease, requires detailed knowledge of the epidemiological parameters related to its infectivity and spread. Systematic data on key epidemiological parameters such as basic reproduction number (a few to several cases to which an individual may pass the disease by direct and indirect contact), incubation period (2 to 14 days and longer when an individual may or may not have symptoms but spreads the disease), sensitivity time (time it takes for a suspected individual to be diagnosed as a confirmed infected case in community), serial transmission time, virus type etc, show large variability (depending upon the socioeconomic, health and environmental factors) and are at early stages of study [1].

Based on such parameters, classical models, like SIR and its variations like SEIQDR [2] and SIDARTHE [3] have been developed for the modelling of COVID-19 disease to guide the implementation of community level interventions to contain and manage the epidemic. However, the parameters related to such models vary, depending upon the ecosystem. These are not easy to assess and fully accommodated in the models. Hence, the accuracy and predictability of the mathematical models for forecasting the epidemic become limiting, though such models are helpful in taking community level decisions.

A powerful alternative approach to capture the temporal components of the epidemic and deploy those for predicting the future course of action is statistical modelling. With the growth of machine learning methods, it has become possible to utilise the statistical principles and build models after learning the principles and parameters from the data itself. With the availability of real time data on the pandemic from nearly 200 countries, it is immediately desirable to apply deep learning methods that learn from country wise data and make predictions, without making the core assumptions required in the epidemiological models. Artificial intelligence (AI) based approaches can handle large data in real time and can utilise global data from the cloud at low costs to make predictions dynamically. Such predictions are highly valuable in guiding policy makers, administration and health departments for data driven management of the pandemic, at levels varying from communities, territories and countries.

A number of research papers have been published in the last six months on the development of models based on multiple linear regression, Gaussian and Bayesian statistics and time series analysis. Some of the recent publications include the application of ARIMA [4] and its variants [4]. The advent of Deep Learning has shown that Recurrent Neural Networks [5] and Long Short-Term Memory (LSTM) [6, 7, 8] outperform previously used models, due to their ability to generalize to diverse series types and provide much higher prediction accuracy. Yet there is need to improve the accuracy and duration of prediction by these models. The AI based models embody the unique feasibility of making quick adjustments in response to any local factors that may impact the course of the epidemic.

This study presents an improved approach to forecast course of the epidemic, compares its performance with other contemporary methods and applies the model to analyze associations between a variety of socioeconomic determinants likely to effect the spread of the infection and the resultant deaths. The prediction model was used to forecast dimensions of the epidemic in 30 countries. A number of socioeconomic predictors that influence infection and death were identified. The model was also applied to predict if the global data on the infections suggests the evolution of COV-2 virus into variants that differed in their degree of virulence. The model was applied to make predictions at desired time points. Once connected to real time data, it can take inputs in real time to re-learn and make autocorrections in short and long term predictions for future dates.

## 2. Methods

### 2.1. Prediction Model

We have earlier reported that the growth of COVID-19 epidemic fits Weibull distribution better than the Gaussian, Beta 4, Fisher-Tippet and Normal Logarithmic distributions [9]. For several countries, the number of daily new infections and deaths was modeled by an improved “Robust Weibull” machine learning approach that fits a Generalized Weibull Distribution (GIW) as shown below:

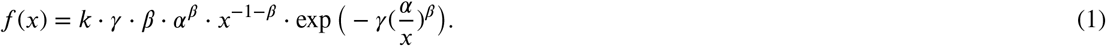

The above model uses an iterative weighting strategy, calculating weights using residuals from the fit curve, to prevent outliers and noisy data that lead to poor curve fits using the Levenberg-Marquardt (LM) method [10] as shown in Algorithm 1. However, this model faces a serious limitation of being very sensitive to new data that may differ even marginally from the data used for building the model. Whenever new data is added, the earlier model is trained from the very beginning. This does not allow predictions to be made with high confidence as the fit curve itself changes significantly with even a single day’s new data. To improve the model in this respect, we developed a time series model to capture the temporal dependence of the model parameters (*k, α, β, γ*) and predict appropriate parameter set.

Series analysis has been used abundantly in the past for various activities like weather forecasting, language translation, etc. Models like ARIMA [4] have prominently been used in the past. However, the advent of Deep Learning has shown that Recurrent Neural Networks [5] and Long Short-Term Memory (LSTM) [11] outperform previously used models mostly due their ability to generalize to diverse series types and provide much higher prediction accuracy.

#### Algorithm 1 Robust Curve Fitting using Iterative weighting

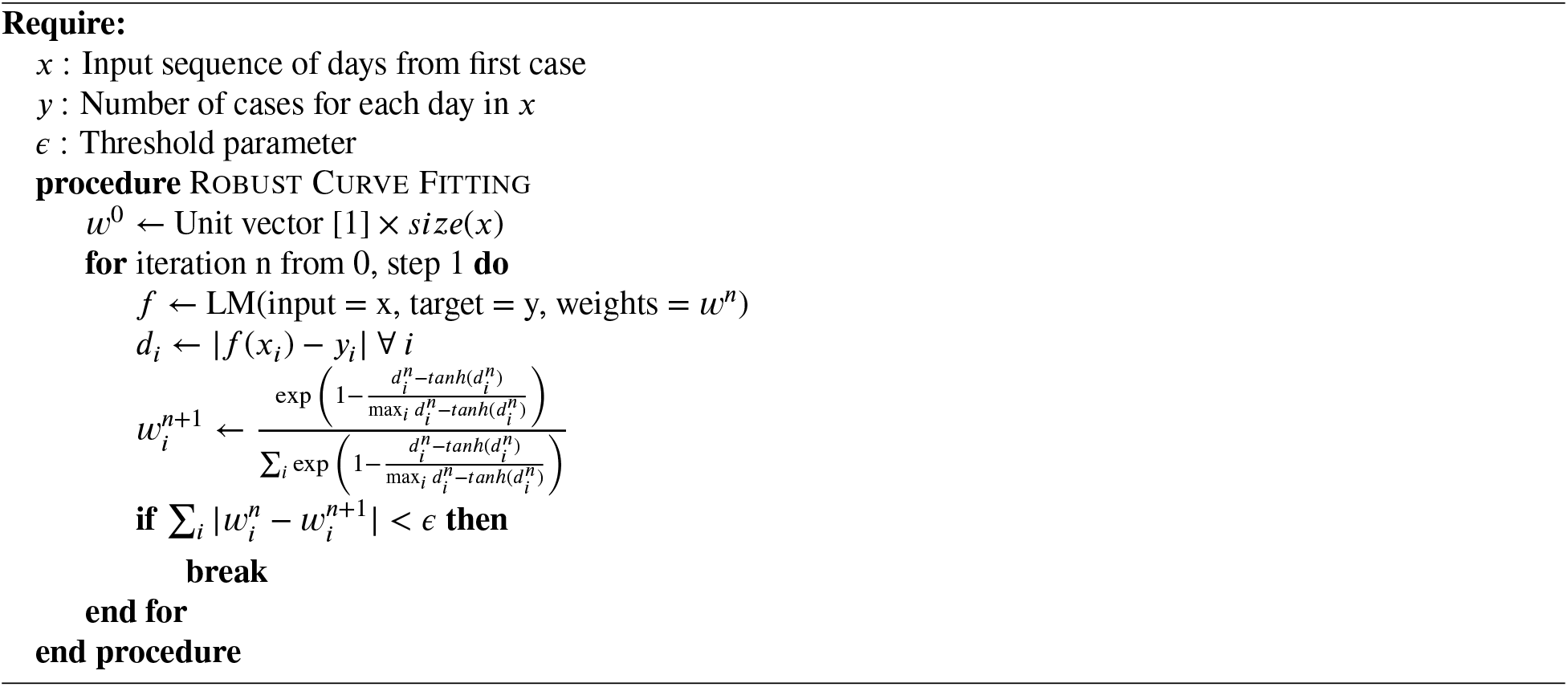

We use LSTM to analyze and predict the best GIW parameters (*k, α, β, γ*). LSTM models use three types of “gates” to analyze the data sequence: input, forget and output gates. A single LSTM cell takes as input the sequence data corresponding to each time-step and a hidden state. Now, for an input at *x*_t_ at time :

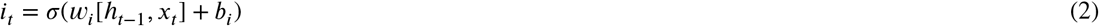

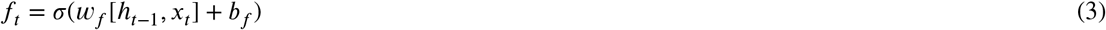

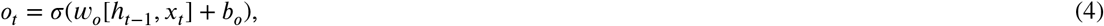

where *i*_*t*_ corresponds to the input gate, *f*_*t*_ forget gate and *o_t_* output gate. Here, each gate has its weight matrix *w* and bias value. Moreover, *h*_*t*− 1_ is the output for the previous time-step. To output *h*_*t*_ and hidden state *c*_*t*_ are calculated as follows:

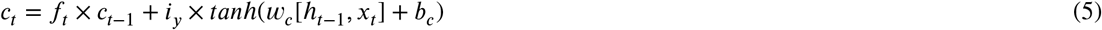

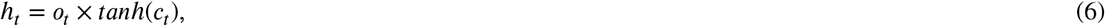

Where *c*_*t* − 1_ is the hidden state of the previous type step. The Equations 2 to 6 can be written compactly as:

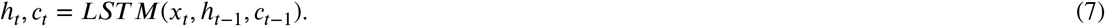

The LSTM model can be applied to a complete time sequence to predict the output at the end of the sequence. Main features of the model are given in Figure 1. The time-series LSTM model is applied on the parameters of the Weibull distribution *k, α, β, γ*. The “iterative” optimization, provided by the LSTM, helps in an under-fitted model to be transformed to a model optimally fitted to the data. The resultant hybrid model is named here as Weibull-LSTM (W-LSTM).

**Figure 1:**
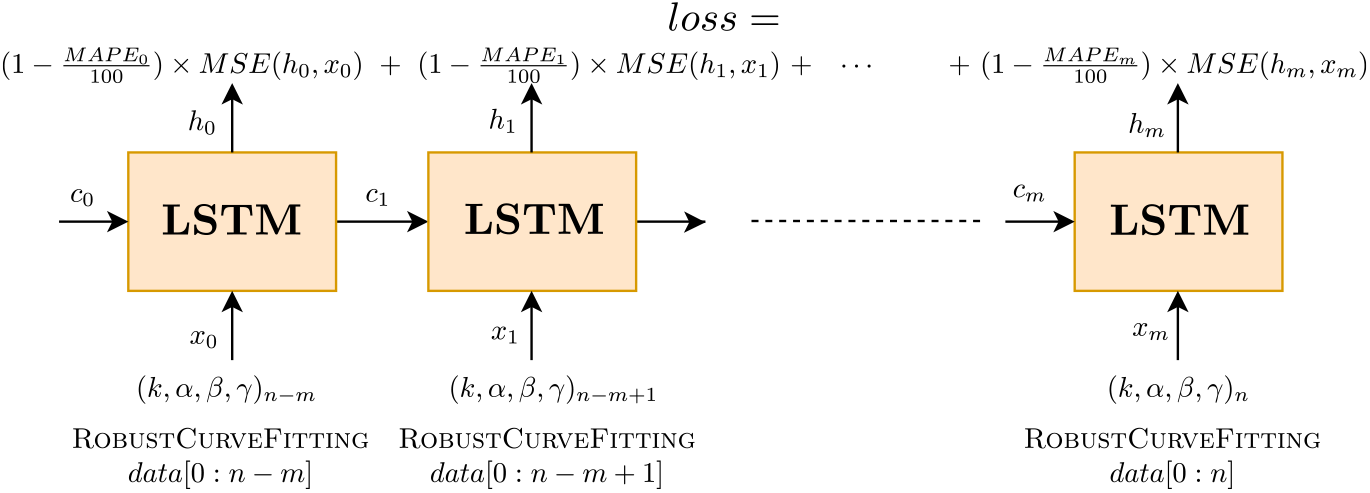
LSTM Model

#### Model Training

To train the W-LSTM model, we use a sequence of 10 curve fits obtained from Robust-Weibull Curve fitting (Algorithm 1). The curve parameters of each of these curves is given as input to the LSTM network in the order of increasing data size as shown in Figure 1. The loss corresponding to each input set is calculated as the Mean Square Error (MSE) between the prediction and the actual *k, α, β, γ* values. This is then weighted by the normalized inverse error 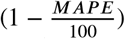, where *MAPE* corresponds to the error of the curve corresponding to the input shape parameters. This gives higher weight to those parameter sets which have lower MAPE scores, allowing the model to converge to better parameter values. Detailed description of the training process is given in Algorithm 2.

##### Algorithm 2 W-LSTM Training

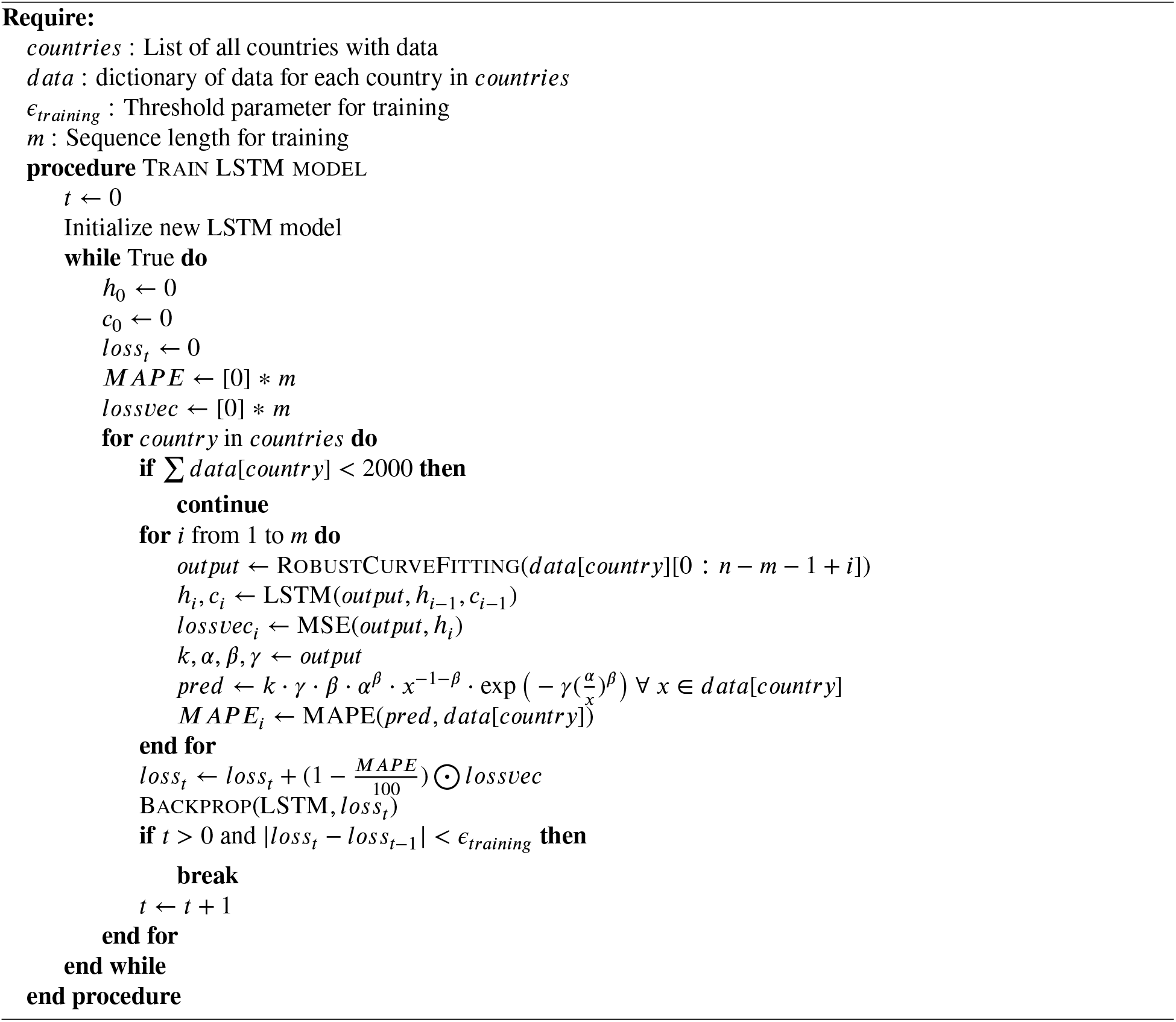

#### Prediction

To predict the best set of parameters for a given data of countries (number of daily new cases/deaths), we use Algorithm 3. Again, the data is used to generate 10 curves. The 10 curves use a subset of original data with last *n* datapoints removed, where *n* decreases from 9 to 0 (*m* in Algorithm 3 is 10 in our case). The LSTM network (trained using Algorithm 2) is given the sequence of curve parameters fit by Algorithm 1 for the 10 curves in a sequence. The final output is used as the parameters of the GIW curve and predictions are made using this distribution.

##### Algorithm 3 W-LSTM Curve Fitting

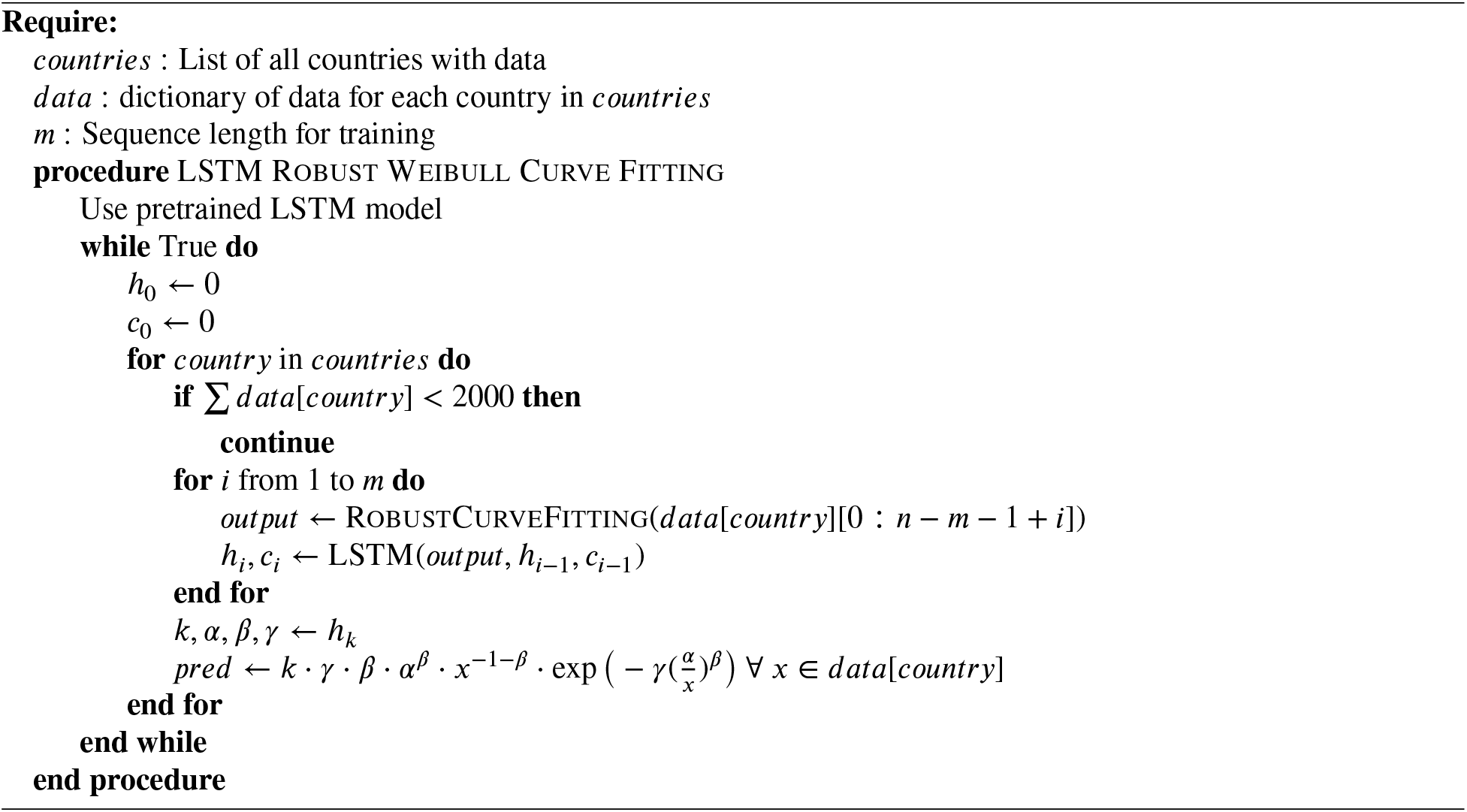

### 2.2. Inferential Statistics

Inferential statistics was applied through Pearson correlation and step wise multiple linear regression analysis. After fitting a regression model, the p-values were used at 5% (p<0.05, highly significant) and 10% (p < 0.1, significant) level of significance to identify the relationships that were statistically significant, and hence most likely influential. To identify the population variables that had highly significant correlation with the disease, only those cases were considered whose r fell beyond the Critical Value (less than the lower limit and more than the upper limit), as applied to two tailed test at n-2 degrees of freedom and 0.05 or 0.1 level of significance. Numerical values of the regression coefficients represent the mean change that the disease (response variable/ dependent variable) will have with one-unit change in the corresponding predictor variable (independent variable), keeping all other factors that influence growth of the disease at constant. Only those independent variables were considered significant whose regression coefficients had p below 0.1, or preferentially below 0.05. Only the variables that were significant at p 0.1 and had a high *R*^2^ were listed as the important predictors of the disease and its severity. The residual plots were examined for unbiased scatter.

### 2.3. Data Sources

#### Epidemic data

The data related to daily new cases of confirmed infections and deaths as a consequence of Covid-19 in different countries was downloaded from Our World in Data COVID-19 Dataset at https://ourworldindata.org/coronavirus. The site updates its data on daily basis from European Centre for Disease Prevention and Control (ECDC). Data till May 19, 2020 was used for learning the model based on W-LSTM. Predictions were made for any desired date, till the numeral value for daily new infections came down to one.

#### Socioeconomic data

The data related to country wise socioeconomic parameters were taken from a number of open source public resources. These include Index Mundi and World Bank at the following links. https://www.indexmundi.com/facts/indicators/SH.MLR.TRET.ZS; and World Bank https://wits.worldbank.org/CountryProfile/en/Country/CHN/Year/2018/.

#### Virus Type data

The data related to the eleven different types of COV-2 virus strains (technically, clades) was taken from https://www.biorxiv.org/content/10.1101/2020.05.04.075911v1.supplementary-material [12] The frequency of the type of COV-2 was determined by the authors in 62 countries, based on the 6181 nucleotide sequences of the genomes sequenced in those countries by April 16, 2020. The frequency data were smoothed by Laplace transform. The original data is given in Supplementary Table 1A. The frequency data before and after smoothing is given in Supplementary Tables 1B and 1C respectively. The Laplace transform data (Supplementary Table 1C) was used in the analysis here.

#### Government Stringency Index

The daily score was calculated as in Oxford Government Response Tracker (https://www.bsg.ox.ac.uk/sites/default/files/Calculation%20and%20presentation%20of%20the%20Stringency%20Index.pdf) and was taken from *Our World In Data*. The score is based on several factors including the closure of schools, market places, works, public gatherings etc. taken as emergency intervention measures to implement social distancing.

## 3. Results

### 3.1. Testing of the W-LSTM model on representative countries

As reported earlier [9] the Robust Weibull curve fitting technique performs superior to other fitting models including - Gaussian, Beta 4, Fisher-Tippet and Log-Normal. In this paper, we have developed an LSTM-based Robust Weibull approach (W-LSTM) and compared with other published methods for the accuracy of prediction based on different estimates of error. We analyzed results obtained from COVID-19 infection data from 5 representative countries : world, India, USA, UK and Italy. Table 1 gives comparison of the fitness accuracy of the W-LSTM model. It gives distinctly better results, as compared to the other distributions, and also the previously used ARIMA [5] and Simple LSTM [6, 7] models that have been applied to COVID-19 infection data. The W-LSTM-based approach developed in this study performs better in terms of MSE, *R*^2^ and MAPE, as compared to the previously published models. This model was therefore applied to several countries for further analysis. The daily new infection and death curves based on W-LSTM model are plotted for 30 countries in Supplementary Figure 1 from the beginning to predicted end of the current epidemic cycle. Curves for the world and few countries with highest cases is reproduced in Figure 2. Data giving analysis of infections, deaths etc. and shape parameters (*k, α, β, γ*) are given in Supplementary Table 2.

**Table 1.** Evaluation of different models on 5 representative countries, for predicting COVID-19 infections. W-LSTM performs significantly better than other distributions. The lowest value of MSE/MAPE and highest values of *R*^2^ are shown in bold.

| Country | MSE | | | | | $R^2$ | | | | | MAPE | | | | |
| --- | --- | --- | --- | --- | --- | --- | --- | --- | --- | --- | --- | --- | --- | --- | --- |
|  | W-LSTM | W-ARIMA | Weibull | Gaussian | LSTM | W-LSTM | W-ARIMA | Weibull | Gaussian | LSTM | W-LSTM | W-ARIMA | Weibull | Gaussian | LSTM |
| World | 3.13E+05 | 3.25E+05 | 3.95E+05 | 4.16E+05 | 3.99E+07 | **0.93** | 0.95 | 0.95 | 0.95 | 0.95 | **39.29** | 40.12 | 49.12 | 42.58 | 60.29 |
| India | 1.93E+01 | 1.96E+01 | 1.98E+01 | 2.08E+01 | 2.19E+03 | **0.91** | 0.96 | 0.94 | 0.94 | 0.93 | **22.00** | 22.13 | 23.44 | 37.03 | 38.44 |
| United States | 1.02E+05 | 1.15E+05 | 1.20E+05 | 1.25E+05 | 1.33E+05 | **0.86** | 0.87 | **0.86** | **0.86** | 0.80 | **26.09** | 104.38 | 27.55 | 134.66 | 149.52 |
| United Kingdom | 9.92E+03 | 9.96E+03 | 9.96E+03 | 1.21E+04 | 1.12E+05 | **0.90** | 0.91 | 0.92 | 0.91 | 0.90 | **14.93** | 14.99 | 15.13 | 104.86 | 93.03 |
| Italy | 2.11E+03 | 2.51E+03 | 2.53E+03 | 5.93E+03 | 3.10E+05 | **0.89** | 0.91 | 0.97 | 0.93 | 0.90 | 12.78 | 12.66 | **12.53** | 214.29 | 58.23 |

**Figure 2:**
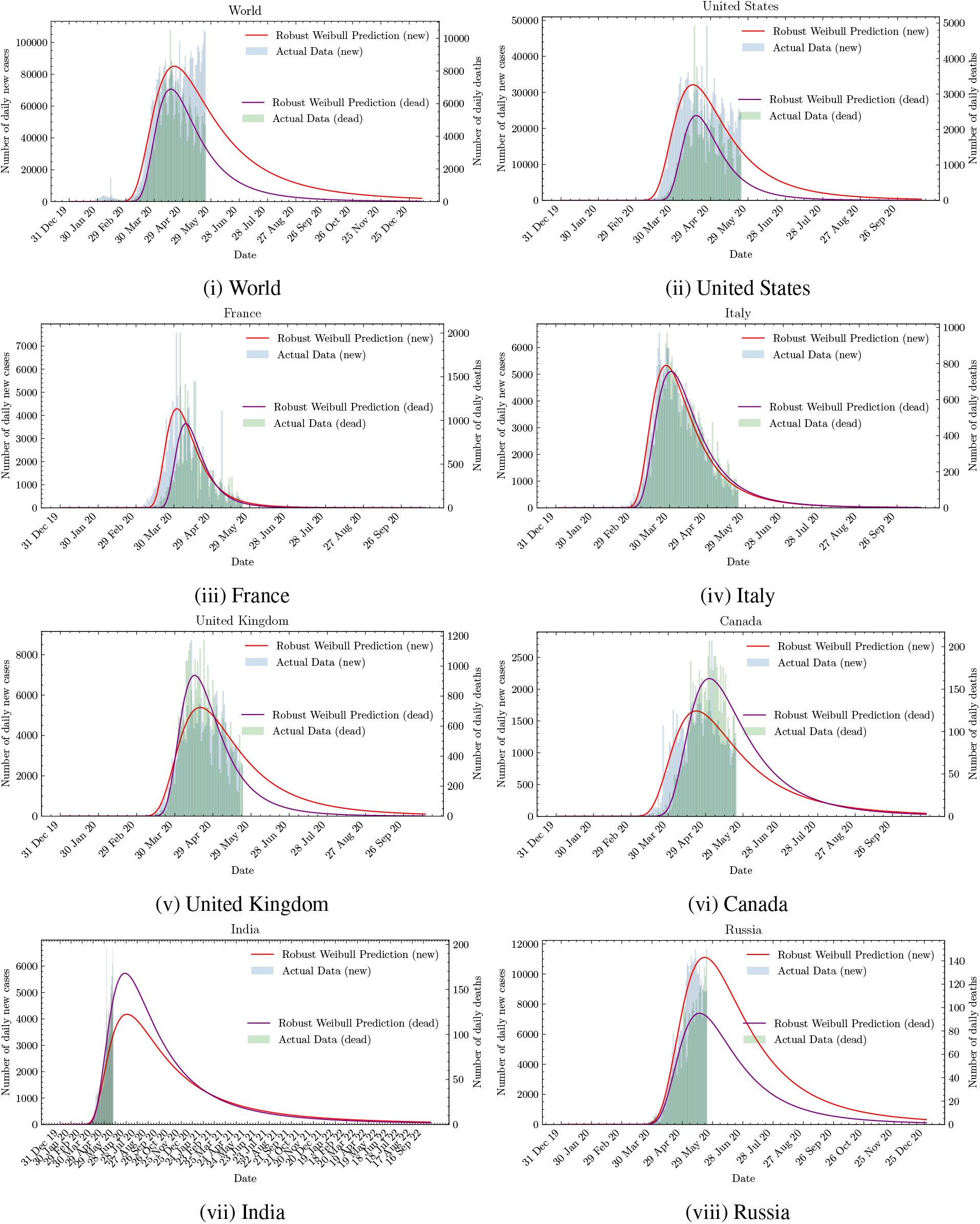
New daily cases and deaths curves for world and a few of the highest affected countries

### 3.2. W-LSTM prediction of infections and deaths till end of the epidemic cycle

The W-LSTM model was applied to publicly available data on daily new cases of infections and deaths from several countries as per the data available till May 19, 2020. Raw data from different countries have lots of missing, incomplete and inconsistent entries, and therefore does not fit any distribution satisfactorily. Out of the 50 countries where sufficient data (for at least 3 months) is available for public view, the W-LSTM model, fits well on 30 countries. These countries were selected for further analysis. Supplementary Figure 1 shows the actual daily values as bars and compares those with the best fitting daily infection and death curves for the predicted values for these 30 countries. Unlike what we had expected, the peak positions of the infection and death curves for the 30 countries, do not show consistency in the survival time indicated by the time difference between the peaks of the curves. This difference should represent the average survival time of population from infection to fatality. In hospital based epidemiological studies, the survival time from disease onset to death varies from 14 to 22 days [13]. As noticed in Supplementary Figure 1, in case of some countries, the deaths data begins and reach the peak height before the infection curve. This is obviously because of late start and large under reporting of infections in several countries, due to insufficient screening of population, unavailability of diagnostic kits in required numbers, inconsistent quality of the diagnostic kits and their high costs [14]. The limitations of diagnostics, along with ambiguities in the interpretation of COVID as the cause of death, differences in average population age and other metabolic disorders etc., also effect the calculations. Due to these reasons, the fatality rates reported from different countries vary from 1 to 7% and more [15]. The survival time difference as indicated by the infection and death curves for the 30 countries varied from -8 to 20 days (Table 2), with an average at 6 days. The small time difference is obviously because in many cases, the infection curve rises slower largely due to under reporting.

**Table 2.**
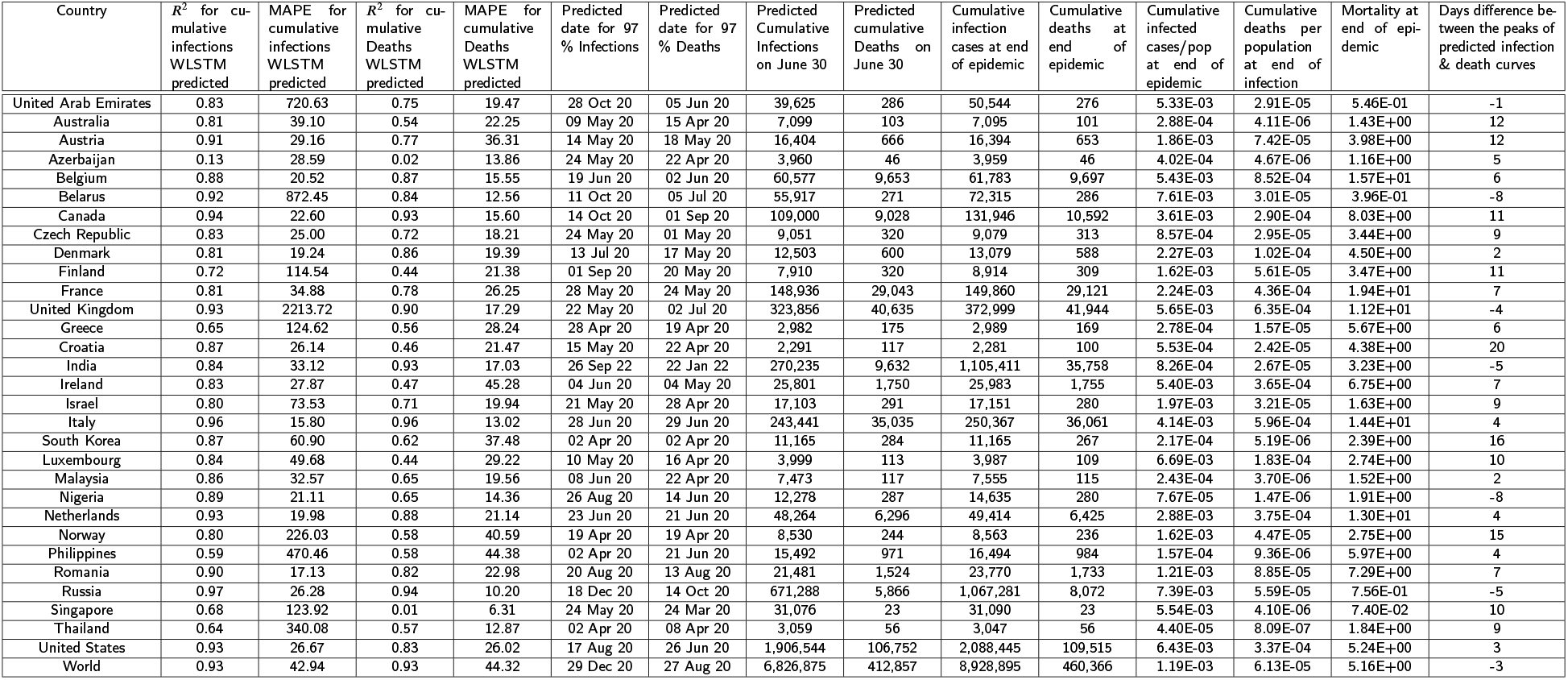
Prediction results for select 30 countries + World.

Table 2 gives the predicted total (cumulative) number of cases for each of the 30 countries as per the W-LSTM model, till the date when the daily new case count is predicted to fall down to single infection. The cumulative number of predicted infections and deaths at end of the epidemic are given in Table 2 along with the country wise dates when 97% of the epidemic will be over. The results show, that out of the 30 countries, India will continue to report cases till as late as the beginning of 2022. The W-LSTM model curve for India (Supplementary Figure 1 and the link http://collaboration.coraltele.com/covid2/) also shows that the country is yet to reach its plateau. As a representative case, Table 2 gives the total number of infections and deaths predicted on June 30th viz., 42 days after the date till when the model was trained.

A validation of the model is given in Supplementary Table 3 where the model trained on data till May 19th was tested for 10 days beyond the period of training on 50 countries. The analysis shows that the predictions by the model trained on data till May 19th matched well with the actual values reported during 10 days following the period of training. The MAPE for the predicted vs actual data on daily deaths to happen during the 10 days after the period of training was less than that for the world (44.32) in 93% of the countries, when the selected 30 countries on which W-LSTM gave good fit were used. At a higher level of stringency viz., MAPE less than 25, the predictions were very good for 70% of the countries (Supplementary Table 3, column C). When additional 20 countries were included, the predictions for daily deaths during the next 10 days beyond the period of training continued to be good. In this case, the MAPE for prediction of deaths during the next 10 days for the 50 countries was less than that for the world in 82% of the countries (Column G). It was less than 25 in case of 58% of the 50 test countries. Quite understandably, the predictions for new infections had higher error rate than those for deaths (Columns B and F), because, as explained earlier, the infections data are highly variable from country to country and have larger proportion of outliers, due to variable approaches to diagnosis of daily new infections.

### 3.3. Identification of likely effectors of infections and deaths

The W-LSTM model was applied on likely factors to have a preliminary analysis of the factors that may influence the growth of the pandemic across countries. To identify such effectors, Pearson correlation was computed between parameters of the epidemic and the likely effectors. The 5 parameters selected for the study were total (cumulative) cases of infection as predicted by W-LSTM till the end of infection cycle, total (cumulative) deaths till the predicted end of cycle, total infections/ population of the country, total deaths/ population of the country and mortality (deaths/infections). The correlations between these 5 parameters and 7 likely effectors are shown in Table 3. Besides the 5 population-based parameters, the likely relationship of the effectors was also estimated in terms of correlation with the 4 parameters (*k, α, β, γ*) related to shape of the Weibull curve.

**Table 3.**
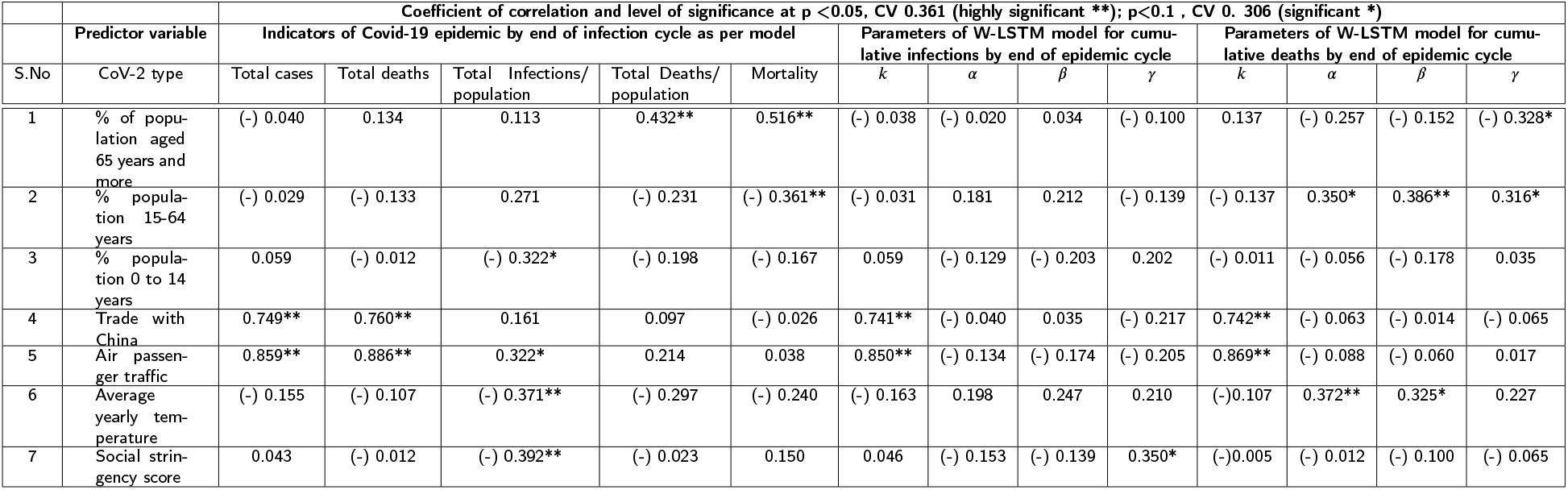
Correlation of COVID-19 infections and deaths (at end of the epidemic as per W-LSTM model) and the parameters (*k, α, β, γ*) of the Model with socio-economic predictor variables. The level of significance of correlation is indicated by ** for p < 0.05 and * for p< 0.1. The significance was determined based on Critical Values (CV) for Pearson Correlation, applied to data on Infections and deaths in 30 countries that gave a good fit in the W-LSTM model (DF 28, p < 0.05, CV is 0.361; p< 0.1 CV is 0.306). The table gives the values of Pearson’s Correlation.

The correlations with *k, α, β and γ* were computed for both the infection and the deaths curves modelled from the beginning of infection to the end of the cycle. As seen in Table 3, a number of highly significant (**) and significant (*) associations are suggested by the correlations. Also, the associations are both in form of promoting the parameter related to the epidemic (positive correlation) and in suppressing the epidemic (Negative correlation). For example, elderly population beyond 65 years in age showed a highly significant positive correlation with mortality. This suggests, the elderly is more likely to get more severe disease and die, as compared to the younger population across the globe. Hence, the countries with more proportion of elderly population are more likely to see deaths. The absence of significant correlation of the above-65 age group with total infections per population suggests that the elderly are likely not more prone to getting infected as compared to others, but are more likely to get a severe disease.

The elderly age group shows a negative correlation (p< 0.1) with the *γ* of death curve, thus making the curve less sharp in rise. In contrast, the adult population (15 to 64 years age) shows highly significant (p < 0.05) negative correlation with mortality, thus suggesting the adults to endure COVID-19 infection and not allow the disease to become severe, hence lower likelihood of leading to death. The endurance of the adult population is reflected in the model by the curve becoming flatter due to higher *α* and *β* (significant positive correlation), besides γ of the death curve. The analysis also shows that the young age group (0 to 14 years) is tolerant to infection by the CoV-2 virus and shows a negative correlation with infection at population level. Hence, the three age groups in population are likely to show different sensitivity and response to different stages of the infection and disease cycle.

Figure 3 illustrates how the lower values of *α* and *β* lead to flatter curves compared to higher values for the constants *k* and *γ*. In this model, *k* increases the peak of the distribution at the same x-coordinate value. On the other hand, increasing *α* and *γ* moves the curve forward and flattens it. However, this behavior is observed on reducing *β*. In that context, *α* and *γ* have opposite effect on the curve shape compared to *β*. If *k* and *γ* are held constant, it is clear from the figure that lower values of *α* and *β* lead to a flattened and elongated curve (purple) compared to the case with higher values of *α* and *β* (red). Thus, a positive correlation with *α* and *β* and a negative correlation with *k* would mean a higher peak with a more prolonged effect of the disease.

**Figure 3:**
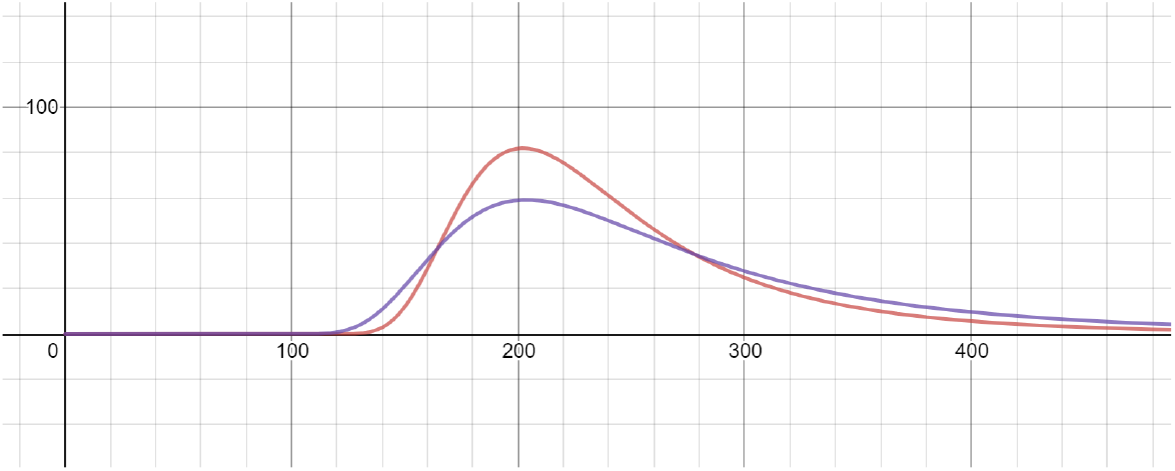
Death cases for constant *k* = 10000 and *γ* = 2500 with (a) higher values of *α* = 38.5, *β* = 4.6 in red and (b) lower values of *α* = 22, *β* = 3.4 in purple.

#### 3.4 Prediction of likely effect of CoV-2 virus type on the pandemic

The various parameters of infection at population level, as predicted by W-LSTM and the parameters related to the shape of the curve were examined for correlation with virus types that have evolved and spread across countries, since initiation of the pandemic in Wuhan in December 2019. The virus types described here, are technically the clades that show differences in the nucleotides in RNA genome of the virus. The SARS-CoV-2 virus has 29926 nucleotide long genome. By now more than 37000 viruses isolated from human patients from different countries have been sequenced across the globe and the sequences deposited at GISAID Database (https://www.gisaid.org). These viruses show about 87% similarity that can be used to cluster them into distinct groups called, clades. Bhattacharya et al. [11] deployed 6181 of the CoV-2 genome sequences and clustered those into 11 groups that are shown in Supplementary Table 1A as the virus types. The frequency of these virus types reported from 61 countries was smoothed by Laplace transform (Supplementary Table 1C) and applied to the group of 30 countries used for modelling the epidemic by W-LSTM. The correlations (Table 4) suggest that the virus type A2a has a highly significant association with deaths / population. Hence this may be the most virulent strain with respect to causing mortality at end of the current cycle of the epidemic. A number of strains show negative correlation with deaths, and thus are infective but less likely killing. Two strains, B and A3 show negative correlation with infections per population, thus suggesting that the countries with prevalence of B and A3 may more likely get a milder disease.

**Table 4.**
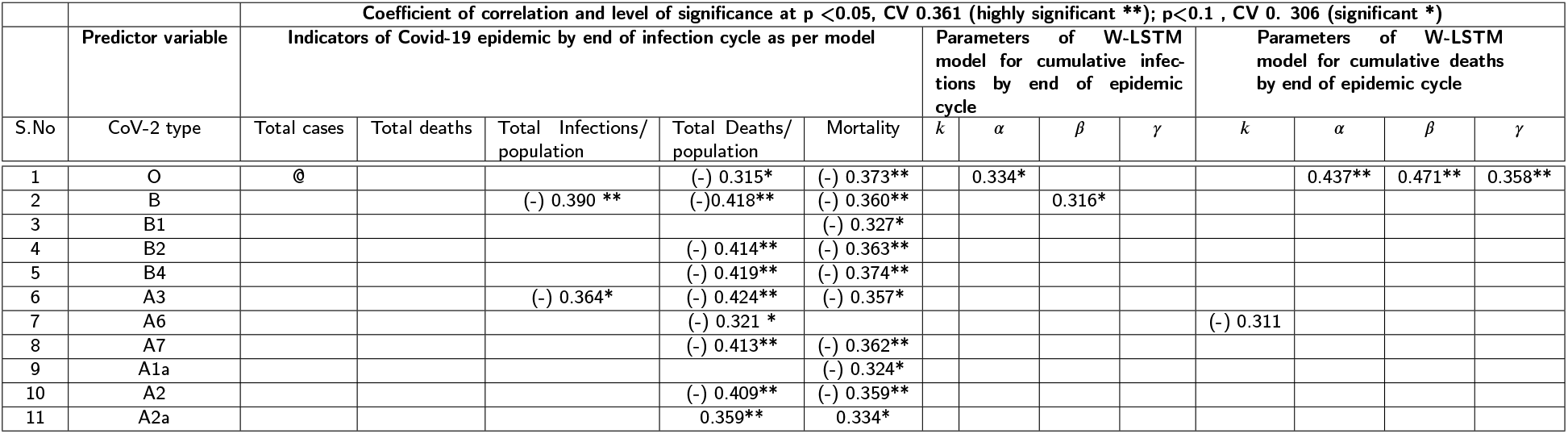
Predicted association of CoV-2 virus types with the WLSTM predicted growth of COVID-19 pandemic across countries. *@: Blank cells indicate the absence of predictions in favor of significant influence*.

## 4. Discussion

The novel coronavirus CoV-2 continues to be leaving a death trail, midst devastating sweep of infection, fear economic loss across the globe. The only strategy to manage the pandemic currently is the non-pharmaceutical approaches. The first need is to have preparedness of the hospitals to manage patients for least distress to patients, quick recovery and minimal mortality. The second important need is to prepare the society for sufficient isolation wards, quarantine centers and social distancing to contain spread of the infection. Identifying infected cases is central to such containment, but non availability and non-affordability of quality diagnostic kits is a serious limitation. Whole population screening is a utopian thought and the savaging character of CoV-2 to continue to be incubated in a large proportion of population without causing any symptoms, weakens all management strategies.

Management of COVID-19 requires robust prediction of the size, duration and dimensions of the ensuing epidemic. As discussed in the background, machine learning methods, backed by good statistical modelling provide a good opportunity to develop on line dynamic systems to prepare the society for managing such epidemic. Because the time series models are based on original numbers from a country, these models may be better predictions of the trend, rather than mathematical models based on epidemiological parameters whose values are often variable, do not account for a variety of socioeconomic, managerial and environmental interventions and involve certain assumptions. The hybrid model developed by us utilizes Weibull distribution as the best fit to the disease data on population and integrates the advantage of self-learning to smooth outliers and train itself in response to shifting trends and fluctuations in data. We have given error based analysis to establish that W-LSTM is superior to other Gaussian and Bayesian distributions used by others. Our model is also superior to other time series applications, including the variations of ARIMA and LSTM. Our analysis shows that W-LSTM trains itself well on country data and makes good predictions with low MAPE for data beyond the period on which the model was trained, as well as for countries that were not part of the constituent data. We have shown that the predictions hold good for 10 days beyond the period of training. We have given predictions for the 42nd day (June 30th, 2020) after the last date of training and for the day when the model predicts the current cycle of epidemic to end. The latter two predictions for future will establish the power and applicability of the model, provided the socioeconomic, environmental and administrative factors remain fairly similar to the conditions as prevailing at the time of the training dataset.

Various population and shape parameters of W-LSTM based modelling of total infections and deaths at end of the epidemic in 30 countries were used to study correlations with a variety of effectors that may influence the spread and severity of the epidemic. Table 5, summarizes a number of predictor variables/ effectors that are likely to exercise significant influence in determining the course of the epidemic. Some of the predictors have promoting influence, while others have suppressing influence. Different age groups clearly respond to CoV-2 infection in different ways. The 65 plus age group is vulnerable to getting the disease in its severe form, though not particularly vulnerable to getting infected. Old age has been suggested as an independent risk factor associated with individuals getting clinically infected with COVID-19 [16]. Meta data analysis suggesting lower vulnerability of children to falling clinically sick with COVID-19 has also been reported [17, 18].

**Table 5.**
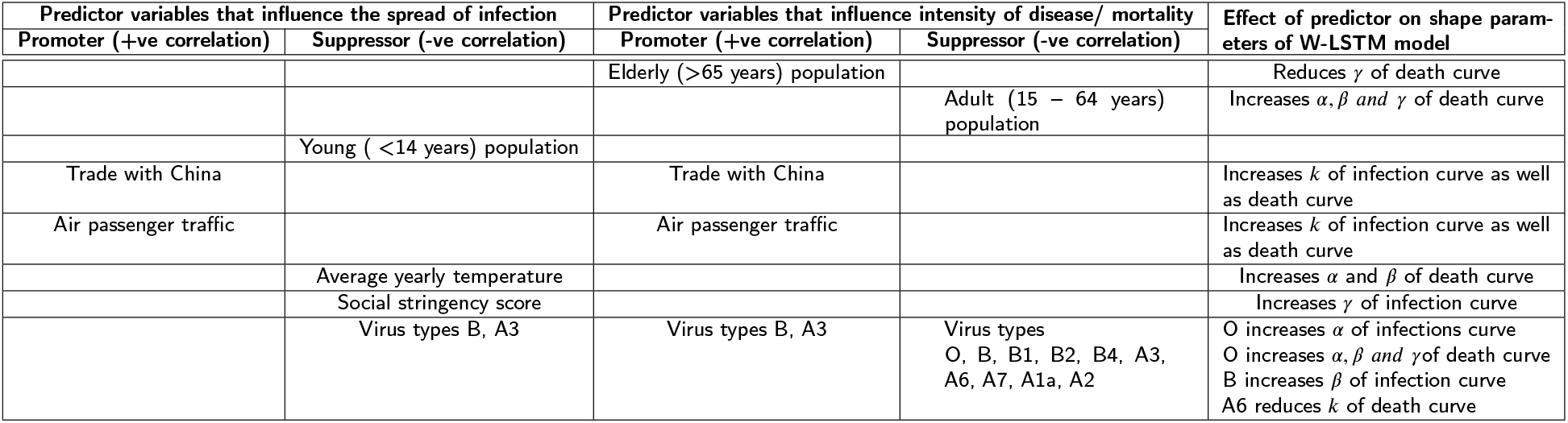
Predictor variables that significantly influence the growth of infection and intensity of disease caused by CoV-2. The inference is drawn from Tables 3 and 4

| Predictor variables that influence the spread of infection |  | Predictor variables that influence intensity of disease/ mortality |  | Effect of predictor on shape parameters of W-LSTM model |
| --- | --- | --- | --- | --- |
| Promoter (+ve correlation) | Suppressor (-ve correlation) | Promoter (+ve correlation) | Suppressor (-ve correlation) |  |
| | | Elderly (>65 years) population | | Reduces $\gamma$ of death curve |
| | | | Adult (15 – 64 years) population | Increases $\alpha, \beta$ and $\gamma$ of death curve |
|  | Young (<14 years) population |  |  |  |
| Trade with China | | Trade with China | | Increases $k$ of infection curve as well as death curve |
| Air passenger traffic | | Air passenger traffic | | Increases $k$ of infection curve as well as death curve |
| | Average yearly temperature | | | Increases $\alpha$ and $\beta$ of death curve |
| | Social stringency score | | | Increases $\gamma$ of infection curve |
| | Virus types B, A3 | Virus types B, A3 | Virus types O, B, B1, B2, B4, A3, A6, A7, A1a, A2 | O increases $\alpha$ of infections curve<br>O increases $\alpha, \beta$ and $\gamma$ of death curve<br>B increases $\beta$ of infection curve<br>A6 reduces $k$ of death curve |

Countries with higher trade with China and higher air passenger traffic show the likelihood of getting higher level of infections. Average yearly temperature in a country, and effective social distancing are likely to have a significant suppressive effect on the spread of COVID-19. Interestingly, the strains of the causative virus CoV-2 show differential association with the growth of infection and severity of the disease. In a recent analysis, high temperature and humidity were reported to reduce the daily new cased and new deaths [19] One strain, namely A2a particularly is likely to be associated with severity of the disease. A number of strains are likely to cause infection but not likely to lead to severity of the disease. Corresponding effect on shape parameters of the model is seen in some cases, as shown in Table 5.

The COVID-19 pandemic is being handled differently in different countries. The government policies related to screening of the population, isolation of confirmed infectious individuals, quarantine of likely infected individuals based on contact tracking, hospital and health care facilities, social distancing measures, socio-economic structure of communities, lock down extent and measures etc., will determine future development of new cases, recoveries and deaths. In response to government measures, the transmission and growth of the epidemic will change dynamically [20].

Currently, a majority of the countries are tapering down closures and lockdowns and are opting for partial lockdown as a strategy to phase out lifting of the lockdown. In some cases, primarily because of unawareness or misinformation, pockets of infected individuals stay unnoticed, and are discovered following a search campaign by the government. In other cases, unplanned migration of workers from cities to rural homelands may shift the territory of infection. Each of these actions affects the distribution of infected and deceased cases in populations differently. This calls for a model to be agile and able to adapt to changing patterns. This can be accommodated in the LSTM-based approach proposed by us. The ARIMA models perform well for stationary time series. LSTMs are beneficial due to the fact that they can store the features of the data for long periods of time. The “iterative” optimization, provided by an LSTM, helps in an under-fitted model to be transformed to a model optimally fitted to the data.

When applying LSTM directly on the number of cases or deaths data (Simple LSTM in Table1), the model is not able to utilize the underlying temporal distribution followed by the data and hence performs poorly. Hence, W-LSTM outperforms approaches trying to model the time-series distribution of cases/deaths directly using LSTM [6, 7]. On the other hand, the W-ARIMA, applies ARIMA on the distribution parameters and only performs well for stationary time series. Further, LSTMs are beneficial due to the fact that they can store the features of the data for long periods of time. The model proposed here can be operated on cloud and linked to global data bases to be updated on real time basis. The model is made to respond appropriately to any temporal changes and learn to modify its forecast. A static version of the model is available on http://collaboration.coraltele.com/covid2/.

## Data Availability

Our prediction model is available online at https://github.com/shreshthtuli/covid-19-prediction. Few interactive graphs can be seen at https://collaboration.coraltele.com/covid2/.

https://ourworldindata.org/coronavirus

https://wits.worldbank.org/CountryProfile/en/Country/CHN/Year/2018/

https://www.indexmundi.com/facts/indicators/SH.MLR.TRET.ZS

https://www.biorxiv.org/content/10.1101/2020.05.04.075911v1.supplementary-material

## Abbreviations

ML: Machine Learning
SARS-CoV-2: Severe Acute Respiratory Syndrome Coronavirus 2
COVID-19: Coronavirus disease

## Acknowledgements

R. Tuli acknowledges the grant provided by Science Engineering Research Board, Department of Science Technology, Government of India towards the JC Bose National Fellowship. Authors acknowledge Coral Telecom Ltd., Noida, India for hosting the model on their server.

## Declaration of Interests

The authors declare that they have no known competing financial interests or personal relationships that could have appeared to influence the work reported in this paper.

---

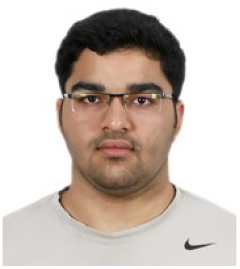

**Shreshth Tuli** is an undergraduate student at the Department of Computer Science and Engineering at Indian Institute of Technology - Delhi, India. He is also a co-founder of Qubit Inc. company which works on providing next generation solutions for industrial problems. He is a national level Kishore Vaigyanic Protsahan Yojana (KVPY) scholarship holder for excellence in science and innovation. He has worked as a visiting researcher at the Cloud Computing and Distributed Systems (CLOUDS) Laboratory, Department of Computing and Information Systems, the University of Melbourne, Australia. His research interests include Internet of Things (IoT), Fog Computing, Network Design, and Artificial Intelligence.

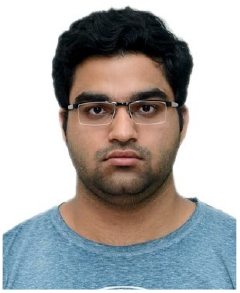

**Shikhar Tuli** is an undergraduate student at the Department of Electrical Engineering at Indian Institute of Technology - Delhi, India. He is the founder and CEO of Qubit Inc. He has worked remotely with the Cloud Computing and Distributed Systems (CLOUDS) Laboratory, Department of Computing and Information Systems, the University of Melbourne, Australia in the realization of the FogBus framework. He has also worked at the Embedded Systems Laboratory, EPFL, Switzerland in the design of low-power and physics-optimized Edge devices made from emerging Non-Volatile Memories. His research interests include Internet of Things (IoT), In-memory and Neuromorphic computing architectures and Nanoelectronics. He specializes in designing novel hardware technologies that are valuable to both industry and academia.

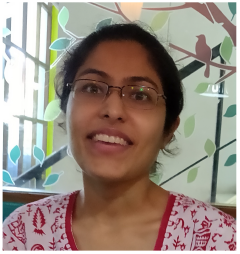

**Ruchi Verma** is an economist with interest in public health and banking. She has deeper interest in socio-economic aspects of human development. She did her Bachelor’ degree from Bombay University and Master’s degree in Economics from University of Sussex, Sussex, UK. She has been working in national and international banks, including HSBC Bank and promoting National Rural Health Mission in India to assist in projects and investments related to socioeconomic and health issues.

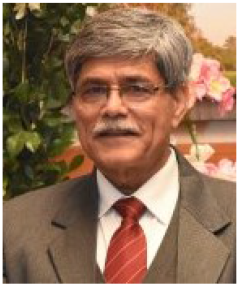

**Rakesh Tuli** is Senior Research Advisor; J C Bose National Fellow, UIET, Panjab University, Chandigarh. Before this, he was executive Director, National Agri-Food Biotech Institute, Mohali; and Director, National Botanical Research Institute, Lucknow. He has many publications in reputed journals and conferences including Nature Biotechnology. His research includes Genomics and Transgenic Approaches to Improving Plants for Agricultural and Health/Medicinal Applications.

## Notes

### Competing Interest Statement

The authors have declared no competing interest.

## References

[1] Marco D’Arienzo and Angela Coniglio. Assessment of the SARS-CoV-2 basic reproduction number, R0, based on the early phase of COVID-19 outbreak in Italy. Biosafety and Health, 2020.

[2] Peng R Zhou C Zhan Y Liu Z et al & merge; Li Y, Wang B. Mathematical Modeling and Epidemic Prediction of COVID-19 and Its Significance to Epidemic Prevention and Control Measures. Ann Infect Dis Epidemiol, 5(1):1052, 2020.

[3] Giulia Giordano, Franco Blanchini, Raffaele Bruno, Patrizio Colaneri, Alessandro Di Filippo, Angela Di Matteo, and Marta Colaneri. Modelling the covid-19 epidemic and implementation of population-wide interventions in italy. Nature Medicine, Apr 2020.

[4] Domenico Benvenuto, Marta Giovanetti, Lazzaro Vassallo, Silvia Angeletti, and Massimo Ciccozzi. Application of the arima model on the covid-2019 epidemic dataset. Data in brief, page 105340, 2020.

[5] Jerome T Connor, R Douglas Martin, and Les E Atlas. Recurrent neural networks and robust time series prediction. IEEE transactions on neural networks, 5(2):240–254, 1994.

[6] Vinay Kumar Reddy Chimmula and Lei Zhang. Time series forecasting of covid-19 transmission in canada using lstm networks. Chaos, Solitons & Fractals, page 109864, 2020.

[7] Anuradha Tomar and Neeraj Gupta. Prediction for the spread of covid-19 in india and effectiveness of preventive measures. Science of The Total Environment, page 138762, 2020.

[8] Ratnabali Pal, Arif Ahmed Sekh, Samarjit Kar, and Dilip K. Prasad. Neural network based country wise risk prediction of covid-19. ArXiv, abs/2004.00959, 2020.

[9] Shreshth Tuli, Shikhar Tuli, Rakesh Tuli, and Sukhpal Singh Gill. Predicting the growth and trend of covid-19 pandemic using machine learning and cloud computing. Internet of Things, pages 100–222, 2020.

[10] Jorge J Moré. The levenberg-marquardt algorithm: implementation and theory. In Numerical analysis, pages 105–116. Springer, 1978.

[11] Sepp Hochreiter and Jürgen Schmidhuber. Long short-term memory. Neural computation, 9(8):1735–1780, 1997.

[12] Chandrika Bhattacharyya, Chitrarpita Das, Arnab Ghosh, Animesh K. Singh, Souvik Mukherjee, Partha P. Majumder, Analabha Basu, and Nidhan K. Biswas. Global spread of sars-cov-2 subtype with spike protein mutation d614g is shaped by human genomic variations that regulate expression of tmprss2 and mx1 genes. bioRxiv, 2020.

[13] Qiurong Ruan, Kun Yang, Wenxia Wang, Lingyu Jiang, and Jianxin Song. Clinical predictors of mortality due to covid-19 based on an analysis of data of 150 patients from wuhan, china. Intensive Care Medicine, 46(5):846–848, May 2020.

[14] Christian G. Daughton. Wastewater surveillance for population-wide covid-19: The present and future. Science of The Total Environment, 736:139631, 2020.

[15] Jean-Louis Vincent and Fabio S Taccone. Understanding pathways to death in patients with covid-19. The Lancet Respiratory Medicine, 8(5):430–432, 2020.

[16] Tao Yu, Shaohang Cai, Zhidan Zheng, Xuejuan Cai, Yuanyuan Liu, Sichun Yin, Jie Peng, and Xuwen Xu. Association between clinical manifestations and prognosis in patients with covid-19. Clinical Therapeutics, 2020.

[17] Nima Rezaei. Covid-19 affects healthy pediatricians more than pediatric patients. Infection Control & Hospital Epidemiology, pages 1–1, 2020.

[18] Tu-Hsuan Chang, Jhong-Lin Wu, and Luan-Yin Chang. Clinical characteristics and diagnostic challenges of pediatric covid-19: A systematic review and meta-analysis. Journal of the Formosan Medical Association, 2020.

[19] Yu Wu, Wenzhan Jing, Jue Liu, Qiuyue Ma, Jie Yuan, Yaping Wang, Min Du, and Min Liu. Effects of temperature and humidity on the daily new cases and new deaths of covid-19 in 166 countries. Science of The Total Environment, 729:139051, 2020.

[20] Melika Lotfi, Michael R. Hamblin, and Nima Rezaei. Covid-19: Transmission, prevention, and potential therapeutic opportunities. Clinica Chimica Acta, 508:254–266, 2020.

